# Using ICD-10-based Social Determinants of Health Categories to Assess Patients Risk for Acute Care Utilization

**DOI:** 10.1101/2021.01.14.21249618

**Authors:** Peter H. Nguyen, James Wang, Pamela Garcia-Filion, Deborah Dominick, Hamed Abbaszadegan, Graciela Gonzalez Hernandez, Karen O’Connor, Shakaib U. Rehman, Sarada S. Panchanathan

**Affiliations:** Department of Biomedical Informatics, College of Medicine, University of Arizona College of Medicine – Phoenix, 475 N 5^th^ St., Phoenix, AZ, United States; Department of Social Work, Phoenix VA Healthcare System, 650 E Indian School Rd, Phoenix, AZ, United States; Department of Clinical Informatics, Phoenix VA Healthcare System, 650 E Indian School Rd, Phoenix, AZ, United States; Department of Biostatistics, Epidemiology, and Informatics, Perelman School of Medicine, University of Pennsylvania, 3400 Civic Center Boulevard, Building 421, Philadelphia, PA, United States

**Keywords:** “Social Determinants of Health (SDoH)”, “Population Health”, “Risk Modeling”, “Electronic Health Records”, “Quality Improvement”

## Abstract

**Background:** There has been an increasing recognition of the influence of social, behavioral, economic, and environmental factors on overall patient health. The purpose of this project was to leverage the ICD-10 codes to identify and link social determinants of health (SDoH) to patients with a high probability of utilizing acute care services and to determine if social service intervention reduced care utilization.

**Methods:** We analyzed retrospective data for active patients at a Department of Veterans Affairs Medical Center (VAMC) from 2015-2017. Eleven categories of SDoH were developed based on existing literature of the social determinants; the relevant ICD-10 codes were divided among these categories. Emergency Room (ER) visits, hospital admissions, and social work visits were determined for each patient in the cohort.

**Results:** In a cohort of 44,401 patients, the presence of ICD-10 codes within the EHR in the 11 SDoH categories was positively correlated with increased acute care utilization. Veterans with at least one SDoH risk factor were 71% (95%CI: 68% - 75%) more likely to use the ED and 71% (95%CI: 65%-77%) more likely to be admitted to the hospital. Utilization decreased with social service interventions.

**Conclusion:** This project demonstrates a potentially meaningful method to capture patient social risk profiles through existing EHR data in the form of ICD-10 codes, which can be used to identify the highest risk patients for intervention with the understanding that not all SDoH codes are uniformly used and some SDoHs may not be captured.

## INTRODUCTION

Social determinants of Health (SDoH) describe several factors that have become growing areas of interest in healthcare due to their impact on patient health and well-being. The World Health Organization define SDoH as “[t]he conditions in which people are born, grow, live, work, and age”(1). While SDoH are commonly thought of only in terms of socioeconomic factors, the term actually broadly encompasses 1) health-related behaviors, 2) socioeconomic factors, and 3) environmental factors (2). There are a variety of factors that comprise SDoH such as tobacco use, diet and exercise, substance use, education, employment, income, family and social support, community safety, and physical environment safety (3).

In 2007, Dr. Steven Schroeder published an article in the New England Journal of Medicine in which he referenced the impact of social and behavioral factors on health and premature death. The combined contribution of social, environmental, and behavioral factors on a patient’s risk for premature death was estimated to be as high as 60%, with genetics contributing 30%, and medical care providing only 10% (4). It is worth noting how low the impact of medical care was estimated to be relative to SDoH.

Ten years after that article, Dr. Sanne Magnan reiterated the importance of SDoH, referencing a model from the University of Wisconsin Population Health Institute’s County Health Rankings & Roadmaps program (2). This model was designed to help identify the factors that impact health in terms of quality of life and longevity in order to stimulate discussion and action in the form of potential changes to policies and programs (5). The County Health Rankings model also illustrated the significant impact of the social determinants on health outcomes, but did not include genetics in its factors influencing health outcomes, as the purpose of this specific model was to look at modifiable factors that could be affected by policy changes. It is interesting to note that, in the County Health Rankings model, modifying health behaviors, social & economic factors, and physical environment could collectively contribute up to 80% towards improving health versus only 20% from modifying clinical care issues. This research again strikingly shows the relatively low impact of direct clinical care, suggesting that SDoH represent a high yield, yet untapped potential for making significant improvements in patient outcomes in healthcare.

The literature has numerous examples describing the impact of different social determinants on health outcomes. Braveman and Gottlieb observed that lower educational status and income were associated with decreased life expectancy at age 25 and increased presence of chronic disease respectively (6). Yelin et al. noted in the literature that poverty appeared to be associated with worse systemic lupus erythematous disease damage (7). Joynt et al. suspected in 2012 that 30-day readmission rates are driven more by a hospital’s patient population and the resources of the community in which it is located than internal factors relating to the hospital itself (8). This appeared to be supported by a recent study by Hu et al. in 2018 demonstrating that a high area deprivation index (ADI) score, which reflects how socioeconomically disadvantaged a geographic region is, was associated with a significantly higher 30-day readmission risk than a lower ADI score (9).

Despite the mounting evidence regarding the role of SDoH on health outcomes, progress in integrating their use has been slow. One of the significant barriers in the past has been in determining a good method to capture the patients’ SDoH profile, though this has become easier in recent years as ICD-10 codes have valid ways to represent many of them to some extent. More recently, the American Medical Association and United Healthcare have collaborated to plan the development of up to two dozen new ICD-10 codes to further identify SDoH risk and integrate it into care (10). The purpose of this project was to leverage the ICD-10 codes to identify and link SDoH to patients with a high probability of utilizing acute care services and to determine if social service intervention reduced care utilization.

## MATERIALS AND METHODS

### Patient population

The data analysis was performed using patient encounter data for an active veteran patient population from January 2017 to December 2017 for a VAMC in a major metropolitan area. The patient cohort was comprised of veterans with an active encounter during two timeframes: in 2017 and from 2015 to 2016. Active encounter data from 2017 were compared to encounter data in the previous two years to examine utilization patterns relative to presence of SDoH and social services intervention. An additional inclusion criterion for this cohort was availability of a means test data within the past five years; the means test includes data on income and employment.

### Exclusion criteria

Test patients and deceased patients were excluded from the dataset. The Charlson Clinical Comorbidity Index (CCCI) of co-morbidities was determined for each patient based on encounter data in 2017 (11). This was used to measure the presence and the degree of major chronic comorbidities (e.g., heart disease, cancer, etc.). Patients with a CCCI greater than zero (i.e., presence of co-morbidities) were excluded from the analysis cohort to reduce bias towards acute care utilization given burden due to major chronic comorbidities.

### Data source

Patient encounter data were extracted from the Regional Clinical Data Warehouse for the VAMC. Microsoft Structured Query Language (SQL) was used for all data querying purposes.

## Identifying Social Determinants of Health Risk

A total of 11 SDoH risk factor categories were determined using previous SDoH work by Schroeder and the Kaiser Family Foundation (3,4). The risk factors were organized by clustering SDoH that were deemed applicable to the VAMC population and that could be readily extracted from the EHR. Schroeder identified that social factors, environmental exposures, and behavioral patterns were modifiable factors influencing health outcomes beyond health care itself. The Kaiser Family Foundation identified 6 major categories which were further subdivided into 29 minor categories. The category of the Health Care System (including coverage and access to a primary care physician) was excluded since the entire study was completed with active patients within a single location within the VA system. The category of food was expanded to include exercise, and categories that have with significant social and medical impact were added (mental health challenges and tobacco, alcohol, and substance use). The final SDoH categories used for this study are listed in Table 1.

**Table 1.** SDoH categories with sample ICD-10 codes and associated social services.

| **SDoH Category** | **Associated Social Services** | **ICD-10 codes** | **Example specific codes** |
| --- | --- | --- | --- |
| Diet and Exercise | Social Work<br>Caregiver Support Services | E66<br>Z71.3, Z71.82, Z72.3, Z72.4 | E66.3 Overweight<br>Z72.3 Lack of physical exercise |
| Employment | Social Work<br>OEF/OIF Support Group (Military-to-veteran transitions)<br>Case Management | Z56 | Z56.0 Unemployment, unspecified<br>Z56.2 Threat of job loss |
| Family/Social Support | Peer Support | Z63 | Z63.0 Problems in relationship with spouse or partner<br>Z63.31 Absence of family member due to military deployment |
| Tobacco Use | • Social Work<br>• SARRTP (Substance Abuse Clinic)<br>• Case Management | • F17.2<br>• Z71.6, Z72.0 | F17.213 Nicotine dependence, cigarettes, in withdrawal<br>Z71.6 Tobacco abuse counseling |
| Education | • Social Work<br>• OEF/OIF Support Group | • Z55 | Z55.0 Illiteracy and low-level literacy<br>Z55.3 Underachievement in school |
| Community Support/Safety | • Peer Support<br>• OEF/OIF Support Group<br>• Community support<br>• Caregiver Support Services | • Z65.1-Z65.3, Z65.8-Z65.9<br>• Z73.6, Z74, Z75.3-Z75.4 | Z65.2 Problems related to release from prison<br>Z75.4 Unavailability and inaccessibility of other helping agencies |
| Alcohol Use | • Social Work<br>• SARRTP<br>• Case Management | • F10<br>• Z71.41 | F10.20 Uncomplicated Alcohol dependence<br>Z71.41 Alcohol abuse counseling and surveillance of alcoholic |
| Income | • Social Work<br>• OEF/OIF Support Group<br>• Case Management | • Z59.5-Z59.9 | Z59.5 Extreme poverty<br>Z59.7 Insufficient social insurance and welfare support |
| Mental Health | • Social Work<br>• Case Management | • F20.3, F20.9<br>• F40.01, F40.10, F40.11, F40.9<br>• F25, F29, F31 - F39 except F34.0 | T14.91 Suicide attempt<br>F20.9 Schizophrenia, unspecified |
| Substance Use | • Social Work<br>• SARRTP<br>• Case Management | • F19<br>• F11 | F11.23 Opioid dependence with withdrawal<br>F19.931 Other psychoactive substance use, unspecified with withdrawal delirium |
| Housing Insecurity | • Social Work<br>• HUDVASH (Homeless Support Services)<br>• Case Management | • Z59.0, Z59.1, Z59.4 | Z59.0 Homelessness<br>Z59.1 Inadequate housing |

When reviewing patient records, one point was assigned per SDoH risk category in which at least one corresponding ICD-10 code was present. A patient could be assigned a total possible cumulative risk score of 11 points if they were found to have at least one ICD-10 corresponding to each category. Multiple codes within the same category were therefore considered to be one entity for the purposes of this work as the emphasis was placed more on how many categories of SDoH were relevant rather than the number of codes per category were present. This would prevent duplication of similar codes from over-estimating the impact of SDoH due to concepts being associated with each other or overlapping in context (i.e. Z65.2 - problems related to release from prison vs. Z65.1 - imprisonment and other incarceration).

### Identifying Acute Care Utilization

Acute care utilization is defined for the purposes of this project as Emergency Room (ER) visits and hospital admissions. The number of ER visits for patients in the cohort was queried with SQL. Hospitalizations were queried by searching for ER disposition fields which corresponded to hospital admissions from the ER since all hospitalizations at this facility are routed through the ER. Both the number of ER visits and the number of resulting hospitalizations were separated by year to correspond to years 2016 and 2017.

### Identifying the Impacts of Social Services Utilization

The impact of social services utilization on acute care utilization was examined by ascertaining social services utilization in 2017 and from 2015 to 2016, and then comparing acute care utilization rates between patients that received any versus no social services utilization. The total amount of utilization by social services were calculated and used in this analysis. Patients were categorized as having either an increase (including newly seen) or decrease (including none) in social service visits in 2017 compared to the prior time period (2015-2016). The patient cohort was further narrowed to patients who had at least one SDoH risk factor based on ICD-10 analysis above and excluded patients who had no acute care services in either 2016 or 2017.

### Data Analysis

The t-test function was used in order to calculate statistical significance. Charts were created with Microsoft Excel, and error bars represent 95% confidence intervals. STATA software was used to perform logistic regression analysis to examine the relationship between individual SDoH risk factors, as well as the total number of SDoH risk factors with ER visits and hospitalizations.

## RESULTS

Based on the initial inclusion criteria parameters, an initial cohort of 59,134 patients was determined. After exclusion criteria stratification, the resulting cohort for January 2017 to December 2017 was 44,401. Of this cohort, 9,379 patients had ER visits and 2,566 patients had hospitalizations during this time interval. Of the total cohort of 44,401, the population had an average age of 58 years +/− 17.4 years and were predominantly male (91%), non-Hispanic (83.2%), and Caucasian (91.8%). In terms of SDoH risk factors in the cohort, 53.3% of patients had at least one risk factor. Details about these demographics and number of SDoH risk factors identified per individual are summarized in Tables 2 and 3, respectively.

**Table 2:** Summary demographics in the acute care utilization cohort.

| **Acute Care Utilization Analysis** |  |
| --- | --- |
| **Cohort Size** | 44401 |
| **Age (Average/StDev)** | 58/17.4 |
| **Gender** |  |
| Male | 91.0% |
| Female | 9.0% |
| **Ethnicity** |  |
| Hispanic | 12.2% |
| Non-Hispanic | 83.2% |
| Unknown / Unreported Ethnicity | 4.6% |
| **Race** |  |
| Caucasian | 81.8% |
| Black | 8.4% |
| Asian | 0.9% |
| American Indian / Alaskan | 1.9% |
| Native Hawaiian or Other Pacific | 0.8% |
| Unknown / Unreported Race | 6.3% |

**Table 3.**
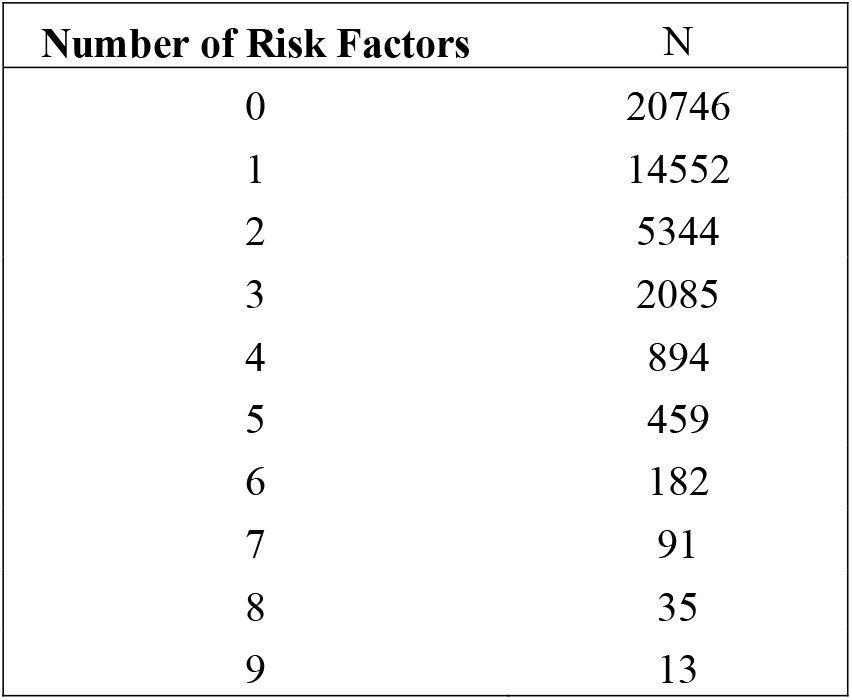
Number of SDoH Risk Factors for patient in acute care utilization cohort.

| **Number of Risk Factors** | **N** |
| --- | --- |
| 0 | 20746 |
| 1 | 14552 |
| 2 | 5344 |
| 3 | 2085 |
| 4 | 894 |
| 5 | 459 |
| 6 | 182 |
| 7 | 91 |
| 8 | 35 |
| 9 | 13 |

Patients who had more SDoH risk factors identified using the ICD-10 categories were likely to have more ER visits and hospitalizations in a given year. Among patients with at least seven SDoH risk factors, the annual ER visit rate was 26 times higher and the annual hospitalization rate was 54 times higher compared to those without a SDoH. The acute care utilization rates by number of SDoH categories is represented in Fig 1.

**Fig 1:**
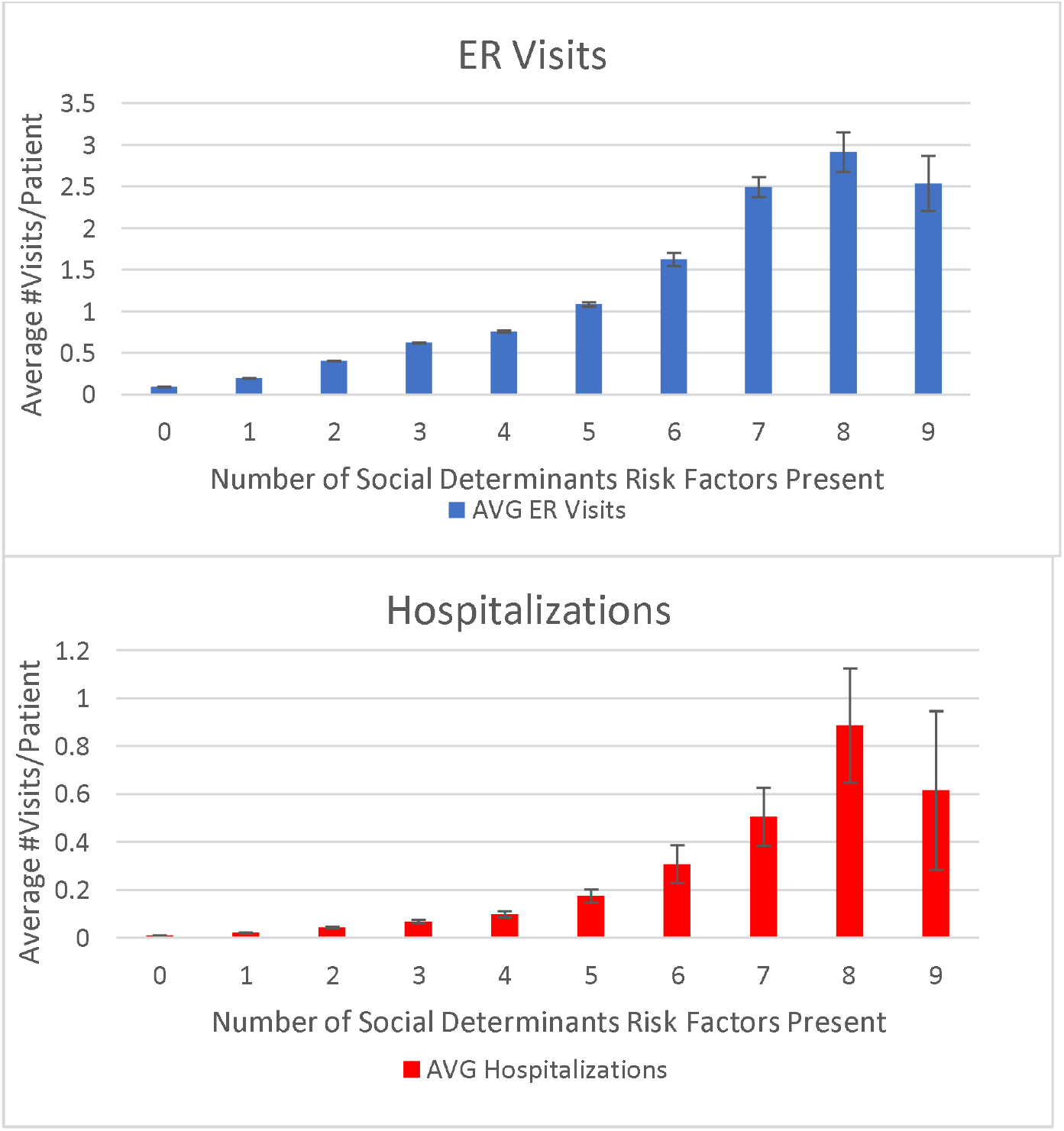
ER visit (a) and hospitalization (b) rates by number of SDoH risk factors for year 2017 for individuals with a Charlson score of zero.

For each SDoH risk factor, the odds ratios (OR) for both ER visits and hospitalization were greater than one and statistically significant, with ORs for each risk factor ranging from 1.77 to 13.01 for ER visits and 1.86 to 18.44 for hospitalizations (Table 4). Patients who had started to use a social service or increased their use of social services had decreased rates of ER visits and hospitalizations, whereas those who had not been seen by social services or decreased their social service utilization had an increase in their annual rate of acute care visits, regardless of social service utilized (Fig 2). Those who used more categories of social services were also more likely to have decreased acute care utilization rates (Fig 3).

**Table 4:** **Logistic regression analysis of the various individual SDoH risk factors, as well as the cumulative total risk score determining their odds ratios on the two outcomes of ER visit or hospital admission.**

| SDoH Risk Factor | Number of individuals with risk factor | Odds Ratio for ER Visits (95% CI) | Odds Ratio for Hospitalizations (95% CI) |
| --- | --- | --- | --- |
| Total | 23256 | 1.71 (1.68, 1.75) | 1.71 (1.65, 1.77) |
| Diet and Exercise | 2892 | 3.19 (2.93, 3.48) | 3.12 (2.62, 3.73) |
| Alcohol Use | 1541 | 3.74 (3.35, 4.18) | 6.15 (5.12, 7.39) |
| Tobacco Use | 2016 | 2.83 (2.56, 3.14) | 3.79 (3.13, 4.59) |
| Drug Use | 437 | 5.74 (4.74, 6.95) | 9.16 (6.98, 12.03) |
| Education | 11 | 13.01 (3.82, 44.6) | 18.44 (4.88, 69.61) |
| Employment | 16620 | 1.77 (1.67, 1.87) | 1.86 (1.63, 2.13) |
| Income | 4786 | 2.27 (2.11, 2.45) | 2.62 (2.24, 3.07) |
| Housing Insecurity | 1476 | 5.41 (4.85, 6.03) | 6.87 (5.73, 8.23) |
| Social Support | 213 | 4.44 (3.36, 5.88) | 3.22 (1.82, 5.66) |
| Community Safety | 2635 | 4.53 (4.16, 4.94) | 6.09 (5.21, 7.11) |
| Mental Health | 6875 | 4.11 (3.86, 4.38) | 4.41 (3.85, 5.05) |

**Fig 2:**
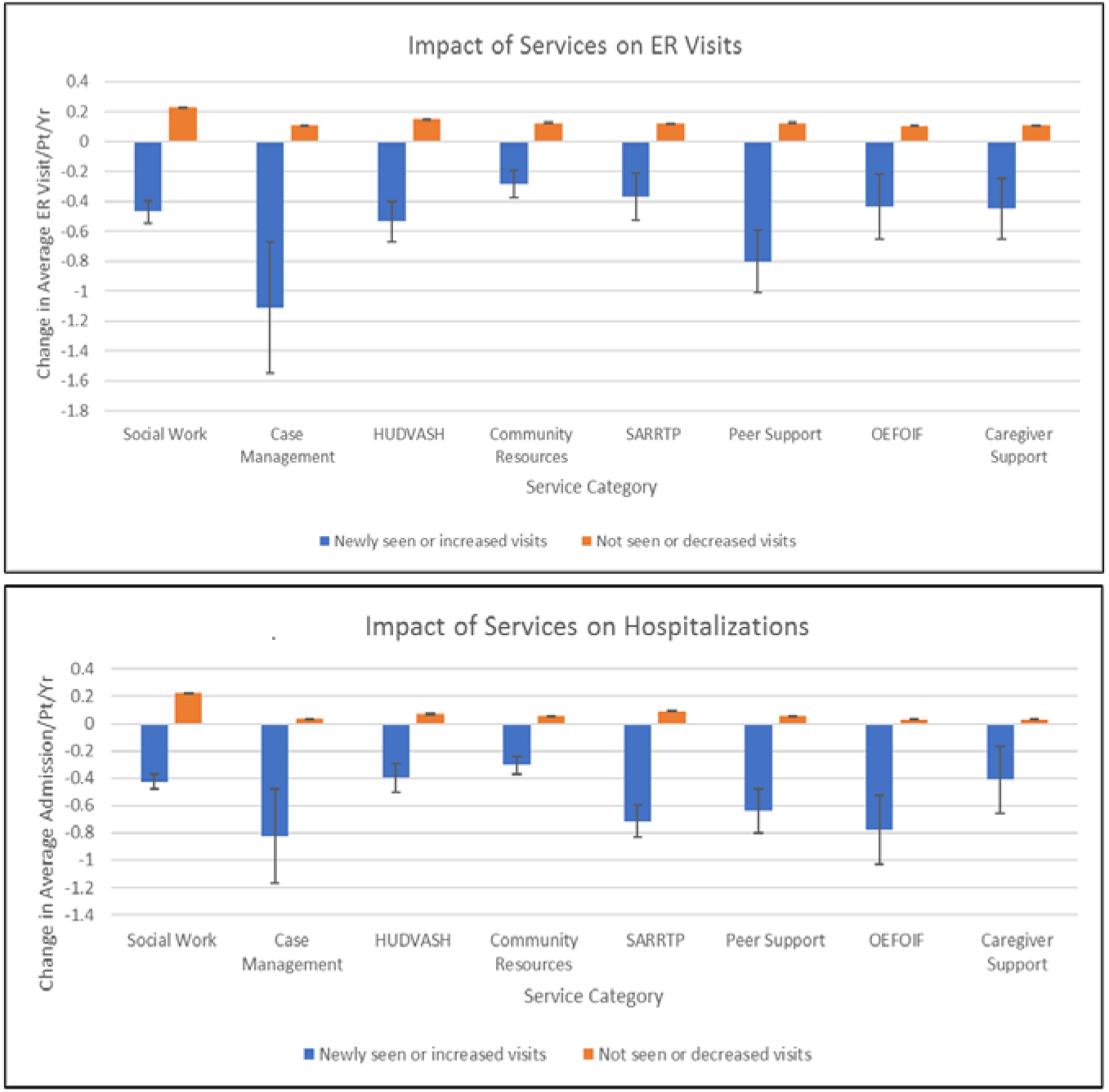
Average annual change in (a) ER visits and (b) hospitalizations in year 2017 compared to 2016 based on whether patients were newly seen (or had increased visits) or were not seen (or had decreased visits).

**Fig 3:**
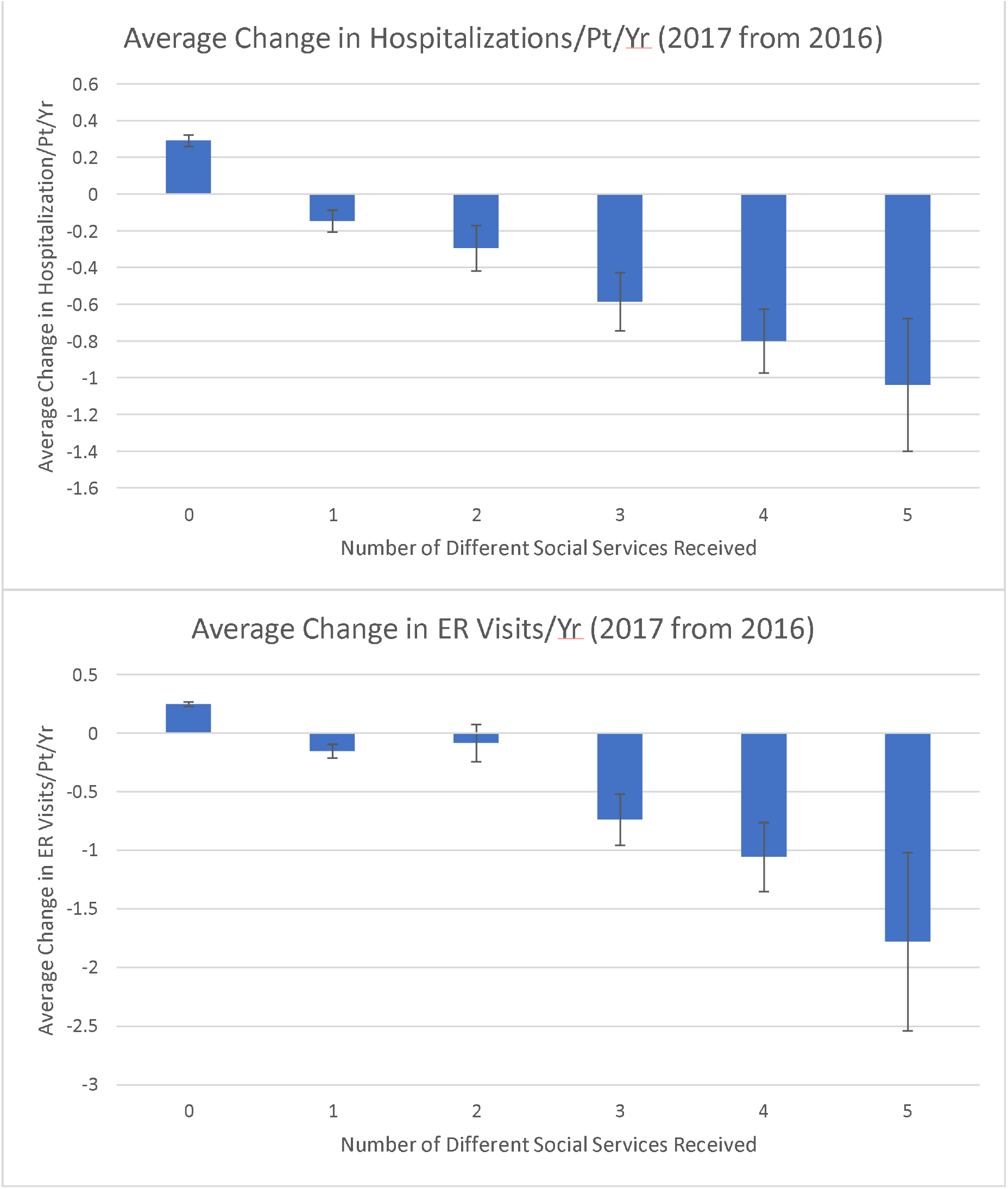
The average change in acute care utilization from year 2016 to year 2017 plotted against the number of distinct Social Services that saw the patient between ER visits (a) and hospitalizations (b) from one year to the next.

## DISCUSSION

Interest in SDoH is increasing rapidly in healthcare and with good reason. As demonstrated by our results and consistent with other studies, ER visits and hospitalizations were occurring at higher rates with increased numbers of SDoH risk factors, which would confirm that there are more factors outside of currently measured healthcare quality factors that influence outcomes and other healthcare metrics (1–9). Having any of the 11 SDoH risk factor categories identified via ICD-10 code in patients without known chronic illnesses was noted to increase the likelihood of acute care utilization; however, certain risk factors appeared to have a more significant impact than others.

On one end of the spectrum, drug use-based and housing insecurity-based risk factors, the categories with higher impacts, had ORs of 5.74 and 5.41, respectively for ER visits and 9.16 and 6.87, respectively for hospitalizations. On the other end of the spectrum, employment-based risk factors, the category with the lowest impact, had ORs of 1.77 and 1.86, respectively. This may indicate what types of risk factors to focus on when planning future interventions and may be helpful in earlier identification of individuals who would be more likely to utilize acute care. Of note, education-based risk factors had ORs of 13.01 for ER visits and 18.44 for hospitalizations. However, in the Veteran population, not only is education not generally coded, all members will have at a minimum a high school diploma or equivalent or they would not have been allowed to enter active duty. Thus, the education category is likely to be very underrepresented in this study (only 11 total individuals with this risk factor) and may not be as reliable in representing SDoH-associated healthcare disparities in the general population.

Social service utilization was found across the board to be positively impactful in reducing acute care utilization, with case management consistently showing the highest level of impact in both the ER visit and hospitalization rates. This makes sense as case management has one of the largest reaches of all the specific social services (seven of 11 SDoH categories impacted) and involves medical care coordination as well. Utilizing more types of social services also decreased rates of acute care utilization likely for a similar reason.

Even with increasing interest in integrating SDoH, the most optimal approach to risk screening is not clear. This project demonstrates that, within the specific healthcare ecosystem of a VA Medical Center, ICD-10 diagnosis codes corresponding to SDoH risk factors, in addition to employment and income data already collected as part of routine VA eligibility means test, can potentially be used to screen for patients for high social risk since ICD-10 has already been integrated into the EHR. Some efforts have been focused on detecting the presence of SDoH mentioned in the unstructured portion of the EHR using natural language processing (NLP) or using machine learning and NLP on a combination of structured and unstructured data (12–21).

### Limitations

While some SDoHs can indeed be ascertained from the EHR, it is important to note that a gap (not quantified in this study) remains: some SDoHs are known, but not coded either due to a lack of a correct code or providers not coding for them. For other SDoHs, there is no adequate code: for example, food insecurity, overcrowded housing, low health literacy or living in an unsafe neighborhood (important reason why people cannot exercise outside). For these, the only recourse is extracting relevant mentions of them in the unstructured portions of the record. In addition, there may be some ICD-10 codes that are missing from this analysis and certain ICD-10 codes could have been allocated to other categories. Despite the limitations, the results of this project remain promising in moving healthcare operations forward to integrate SDoH and mitigate its negative effects.

The optimal method of capturing SDoH remains debatable and some factors may not necessarily be captured by the method in this project. For example, legal troubles, cultural norms and barriers, and suspicion about the healthcare system could be perceived as social risk factors, however they would not neatly fall into the risk factor categories used in this project.

Furthermore, the identification of risk factors in this project is highly dependent on optimal data entry by clinicians and healthcare employees. If clinicians do not document ICD-10 codes for a patient with housing insecurity, level of education or issues with family support, this method will only sub-optimally describe that patient’s social risk profile. This limitation should not be understated, particularly given the current challenge for healthcare workers to accurately document encounters in the setting of increasing documentation requirements. For instance, the high OR of acute care utilization for the education SDoH risk factor with a wide confidence interval may be due to artifact from limited data availability. Also, specific to the VA system, some data elements such as income level may be more readily available than in other healthcare systems due to the nature of how VA benefits are determined. In addition, the cohort in this project was predominantly male, Caucasian, and non-Hispanic and is not representative of the general patient population in the United States.

Additionally, it should be stated that in the analysis of the utilization of social services, patients merely having an encounter with a social service provider is being interpreted as a surrogate for receiving some kind of social service benefit, such as resources, food, or housing. However, it may be the case that having an encounter with a social service might result in no assistance being available for a patient. Ultimately, it should be noted that the results of this project identify correlations, not necessarily causations.

## CONCLUSION

Given the increasing importance of the SDoH and their integration into care, the findings from this quality improvement project are important because they represent an efficient method in which SDoH can be captured and linked to health outcomes. Most EHR systems do not yet incorporate SDoH risk screening as part of their data collection, nor do many clinical workflows. The use of the ICD-10 codes does not require any new modification to EHRs.

Furthermore, the impact of utilization by the various social services on acute care utilization is encouraging and suggests a means to make SDoH data actionable. Future work will revolve around operationalizing these findings through personalized healthcare and potential use of new modalities such mobile medical applications as well as improved methods of data capture such as NLP.

## Data Availability

Data not available due to ethical/legal/commercial restrictions. Due to the nature of this research, participants of this study did not agree for their data to be shared publicly, so supporting data is not available.

## Acknowledgements

We would like to thank Amit Dahiya, Michael Smith, Don Copp, Pete Fredericks, and Daniel Garcia of the Clinical Informatics Analytics and Applications Department at the Phoenix VA Medical Center for their support with SQL, guidance on clinical data warehouse navigation, and data analysis support. We also acknowledge Dr. Umar Iqbal, Dr. Behnam Vahdati Nia, Dr. Ali Al-Yaqoobi, and Dr. Vikeen Patel from the University of Arizona College of Medicine – Phoenix for their input and support during the execution of this project.

